# Gene expression levels of the glycolytic enzymes lactate dehydrogenase A (LDHA) and phosphofructokinase platelet (PFKP) are good predictors of survival time, recurrence and risk of death in cervical cancer

**DOI:** 10.1101/2022.08.02.22277946

**Authors:** Verónica Bolaños-Suárez, Ana Alfaro, Ana María Espinosa, Ingrid Medina-Martínez, Eligia Juárez, Nicolás Villegas-Sepúlveda, Marco Gudiño-Zayas, América Gutiérrez-Castro, Edgar Román-Bassaure, María Eugenia Salinas-Nieves, Sergio Bruno-Muñoz, Oscar Flores-Herrera, Jaime Berumen

## Abstract

**Background:** Up to 74% of patients with cervical cancer (CC) may experience recurrence after their treatment, and most of them are identified late when only the clinical parameters are used, which decreases their chances of recovery. Molecular markers can improve the prediction of clinical outcome and identify therapeutic targets in CC. Glycolysis is altered in 70% of CCs, so it could be a metabolic pathway in which molecular markers associated with the aggressiveness of CC can be identified.

**Methods:** The expression of 14 glycolytic genes was analyzed in 118 CC samples by microarrays, and only LDHA and PFKP were validated by qRT–PCR (n=58) and in second and third replicates by Western blotting (n=69) and immunohistochemistry (n=18).

**Results:** LDHA and PFKP were associated with poor overall survival [OS: LDHA HR=3.0 (95% CI= 1.1-8.2); *p=2*.*9 × 10*^*-2*^; PFKP HR=3.4 (95% CI= 1.1-10.5); *p= 3*.*5 × 10*^*-2*^] and disease-free survival [DFS: LDHA HR=2.7 (95% CI= 1.6-6.3); *p=2*.*6 × 10*^*-2*^] independent of FIGO clinical stage. The risk of death was greater when both biomarkers were overexpressed than when using only FIGO stage [HR =7 (95% CI 1.6-31.1, *p=1*.*0 × 10*^*-2*^) versus HR=8.1 (95% CI=2.6-26.1; *p=4*.*3 × 10*^*-4*^)] and increased exponentially as the expression of LDHA and PFKP increased.

**Conclusions:** LDHA and PFKP at the mRNA and protein levels were associated with poor overall survival, disease-free survival and increased risk of death of patients with CC regardless of FIGO stage. The measurement of expression of these two markers could be very useful to evaluate the clinical evolution and the risk of death from CC and to make better therapeutic decisions at the beginning of treatment.

## 1. Background

Cervical cancer (CC) is the fourth most common cancer in women worldwide [1] Persistent infection with human papillomavirus (HPV), particularly high-risk oncogenic viruses, is the main etiological agent for the development of CC. Despite early detection programs and vaccinations against the most oncogenic HPVs [2], it is estimated that 569,000 new cases occur each year and cause 311,365 deaths worldwide [1], indicating that CC continues to be a major health problem, mainly in developing countries where most cases occur.

Treatment of CC includes surgery, radiotherapy and chemotherapy, which depend on the clinical stage of the disease [3]. However, it is estimated that a significant percentage of patients have pelvic recurrence (10-23% in stage IB-IIB and 42-74% in stage III-IVA) or metastases (16-26% in stage IB-IIB and 39-75% in stage III-IVA) after treatment, and when this occurs, the patients often have poor survival [4], [5]. In addition, only 32% of patients with recurrent disease are identified early (before 6 months) during medical follow-up, which decreases the chance of recovery and the survival time [6]. Although the clinical International Federation of Gynecology and Obstetrics (FIGO) stage, the clinical characteristics of the tumor, metastasis to lymph nodes and parametrial invasion are predictors of recurrence and survival time, there are no molecular markers approved for clinical use that predict the clinical evolution of patients with CC. Molecular markers alone or in conjunction with clinical data improve the prediction of the clinical outcome and facilitate better therapeutic decision-making, as has been demonstrated in colorectal [7] and breast [8] cancers.

Since glycolysis is increased in 70% of human cancers, with lactate production even in the presence of oxygen (Warburg effect) [9][10], it can be used to identify new prognostic biomarkers in CC. In fact, the usefulness of several genes or proteins involved in glycolysis has been investigated to evaluate survival and aggressiveness in CC, such as glucose transporter 1 (GLUT1) [11], hexokinase 2 (HK2) [12], phosphofructokinase isoform M (PFKM) (as part of a genetic profile)[13] and total lesion glycolysis (TLG), a parameter measurable through positron emission tomography (PET) [14]. However, the predictive efficacy of these biomarkers either has not been reported or has been reported to be intermediate. Perhaps PET is the most efficient method; however, it is very expensive. On the other hand, it would be desirable to identify molecular therapeutic targets in CC since the antitumor strategy with specific target drugs is showing great benefit in many tumor types with a marked decrease in side effects [15].

In a previous study, our group found a glycolytic gene profile in CC associated with a decrease in survival [16]. In this paper, we investigate which genes of that glycolytic expression profile are most highly associated with survival and tumor aggression in CC. Of the genes finally identified, the mRNA and protein levels of lactate dehydrogenase A (LDHA) and phosphofructokinase platelet (PFKP) were explored in a main discovery sample of 118 CCs, and they were validated in two additional replicates (n = 58 and 69 CCs) and in 18 CC tissues preserved in paraffin. These two markers allow to evaluate of the clinical evolution and mortality of patients with CC regardless of the FIGO stage.

## 2. Materials

### Ethical consent

The study protocol was approved by the Scientific and Ethics Committee of the General Hospital of Mexico (HGM) (approval number DIC/03/311/04/051). This study, all experiments and analyses were performed in accordance with the Declaration of Helsinki [17]. Written informed consent was obtained from all participants before their inclusion in the study.

### Patient selection and clinical characteristics

The study included 188 patients with CC (Supplementary Figure 1, Supplementary Table 1), 10 patients with high-grade epithelial lesions (HG-CINs) and 36 women with healthy cervical epithelium (HCE) evaluated in the Departments of Oncology and Gynecology and Obstetrics of the HGM. Patients with CC were selected from a previous study that included 462 patients recruited from November 2003 to April 2005 and from January 2006 to July 2007 [18]. The inclusion criteria were as follows: patients diagnosed with invasive CC, without previous treatments. Only patients from whom high-quality RNA and tumor biopsies had more than 70% tumor cells were included in the present study. The exclusion criteria were insufficient quality of the biological sample. All patients received complete clinical evaluation and were treated with surgery, radiation, chemotherapy, or a combination of these according to American Cancer Society guidelines. Tumor staging was performed in accordance with the latest International Federation of Gynecology and Obstetrics (FIGO) protocol for gynecological cancer [19]. The average age of the patients was 51 years, with a range of 23 to 89 years. After their treatment, they were followed up and evaluated periodically at the HGM. HCE were obtained from patients who underwent hysterectomy for myomatosis with a normal cervix for cytology and colposcopy as described previously [18]. The tissues were frozen in liquid nitrogen and stored at - 80°C.

### RNA isolation

Total RNA from the samples was extracted with TRIzol™ reagent (Invitrogen, Carlsbad CA, USA) according to the manufacturer’s instructions. RNA integrity was verified by agarose gel electrophoresis, determining the presence of 18S and 28S ribosomal RNA.

### Glycolytic gene expression and data analysis

The glycolysis gene expression of 76 CC, 10 HG-CIN and 17 HCE tissues was examined using a Human Gene 1.0ST (HG-1.0ST) microarray and with the Human Gene Focus (HG-Focus) microarray in 42 CCs and 12 HCE (Affymetrix, Santa Clara, CA). Gene expression was obtained from two studies previously published by our working group in the Gene Expression Omnibus (GEO) database with accession numbers GSE52904 [16] and GSE39001 [18]. See supplementary text 1.

### Quantitative PCR (qRT–PCR)

cDNA was synthesized using the High-Capacity cDNA Transcription Kit (Applied Biosystems) using 5 μg of RNA according to the manufacturer’s protocol. Gene expression of *LDHA, PFKP* and an internal control (*RPS13*) was measured in 58 CC and 19 HCE by qRT–PCR and TaqMan gene expression assays were used (*LDHA*, Hs00855332_g1; *PFKP*, Hs00242993_m1; RPS13, HS 01011487_g1; Applied Biosystem Inc.). The experiments were run in triplicate in a final volume of 20 μL, including 200 ng of cDNA template using the TaqMan® Universal PCR Master Mix (4304437, Applied Biosystems), according to the manufacturer’s instructions. The expression of the LDHA and PFKP genes was normalized in each tumor and control sample with *RPS13* using as previously reported [18]. The FC in expression was calculated by dividing the median normalized intensity of each tumor sample by the median normalized intensity of the whole control samples.

### Western blot (WB)

LDHA and PFKP protein expression was determined using WB in 69 CC samples. 25 ng of protein was resolved on 8% SDS-PAGE and electrotransferred onto a nitrocellulose membrane and incubated with a mouse monoclonal antibodies anti-human LDHA (H-10: sc-133123; 1:1,000) or PFKP (F-7: sc-514824; 1:200) and goat β-actin antibody (I-19: sc-1616), (Santa Cruz Biotechnology, Inc.) overnight at 4°C. The membranes were then incubated with horseradish peroxidase (HRP)-conjugated secondary antibodies (anti-mouse IgG + IgM (H+L) antibodies; 1:10,00; Jackson ImmunoResearch Laboratories, Inc.) and anti-goat IgG-HRP (sc-2354; 1:1,000; Santa Cruz Biotechnology, Inc.) for 1 h at room temperature. Prestained Broad Range SDS–PAGE Standards (BIO-RAD, CA) were used for molecular weight estimation on gels. β-actin was used as an internal control. Loading buffer without sample was used as negative control. The immunoreactive proteins were developed using the SuperSignal™ Chemiluminescent HRP Kit (Thermo Fisher Scientific). Densitometric analysis was performed with ImageJ (NIH, Bethesda, MD). The optical density was calculated as OD=Log 10 (255/pixel value).

### Immunohistochemistry (IH)

The protein expression of LDHA and PFKP was determined in 18 CC, 12 HCE and 6 metastatic samples by IH. Human paraffin-embedded tissue samples were collected at the Pathology Department of HGM from patients evaluated from January 2008 to March 2013. The inclusion criteria were women with CC at any FIGO stage whose diagnostic biopsy was taken prior to treatment, with complete clinical data available, and with follow-up data of at least 24 months after treatment. All patients received complete clinical evaluation according to the ACS guidelines. Clinicopathological information was collected from medical records. Tissue microarrays (TMAs) were built as previously described [18]. The TMA included kidney tumor tissue as a positive control, which was previously determined to present high LDHA and PFKP expression (www.proteinatlas.org). IH was performed with the Ultra Streptavidin (USA) HRP Detection Kit (Multi-Species) (BioLegend, CA) according to the manufacturer’s instructions. The following mouse monoclonal antibodies were used: LDH (H-10) sc-133123 (1:200) and PFKP (F-7) sc-514824 (1:100) from Santa Cruz Biotechnology (Santa Cruz, CA). Antigen-antibody complexes were detected using the avidin-biotin peroxidase method, with 3,3′-diaminobenzidine-tetrahydrochloride as a chromogenic substrate (DAB Chromogen Concentrate, BioLegend, CA), and the sections were counterstained with hematoxylin. Assays were performed in triplicate.

### Quantitative image analysis

Each tissue of the TMA was photographed in triplicate with a magnification of 400X using a Nikon Microphot FXA. The digital images were analyzed with ImageJ as previously described [20] and the intensity of DAB signal was transformed to optical density values: OD= -log (255* maximum level pixels)/average pixels. The integrated optical density (DOI) was calculated as OD x staining area.

### Survival analysis

After the treatment was completed, each patient was clinically evaluated every 3 or 6 months by an experienced oncologist. Clinical data of the follow-up study were obtained from the patient’s medical record. Additionally, a social worker performed phone calls and home visits to the patients every 6 months during the study. Survival analysis was performed on all patients who received the full treatment. The mean follow-up time was 60 months after the initial diagnosis. The patients designated as censored referred to patients who were lost to follow-up or who died from causes other than CC. Patients were considered lost when they did not attend medical appointments for disease control, were not found at home visits or did not answer phone calls. In this cohort, alive status was registered at the last follow-up, and death was caused by a primary tumor of CC as a main cause and confirmed by the medical record and the death certificate.

### Statistical analysis

Data analysis was performed using SPSS software ver. 20. Receiver operating characteristic (ROC) curve analysis was performed, and the Youden index was used [21] to select the best cutoff points to distinguish tumors from nonsurviving and surviving patients or patients who survived with and without the disease using the expression values of glycolysis genes obtained by microarrays, qRT–PCR and WB. Expression values equal to or above the cutoff were considered upregulated, and those with values below the cutoff were considered downregulated.

The comparisons of the overall survival (OS) and disease-free survival (DFS) times between patients with upregulated and downregulated tumor genes were calculated by the Kaplan–Meier method and analyzed by the log-rank test. FIGO staging and glycolysis gene expression were included in univariate and multivariable Cox proportional hazards regression models. All tests were 2-sided, and *p* values less than 0.05 were considered statistically significant.

## 3. Results

### Analysis of glycolytic gene expression in CC

The expression of genes involved in glycolysis included in the HG-1.0ST microarray was analyzed in 103 cervical samples, including 76 CC, 10 HG-CINs and 17 HCE. As shown in Supplementary Figure 2, the 14 genes had significantly higher expression levels in the invading CCs than in the controls. Interestingly, we were able to confirm the difference between the invading cancers and the control group for 9 of these genes (SLC2A1, HK2, PFKP, ALDOA, GAPDH, PGK1, ENO, PKM and LDHA) in a second replicate with 42 invasive CCs and 12 controls, explored with a second microarray (HG-Focus; see Supplementary Figure 3), which included only 9 of the 14 formerly explored genes. The samples were distributed into 3 groups, according to the expression profile, in the hierarchical clustering analysis: upregulation (group 3); intermediate regulation (group 2), which could be divided into the intermediate downregulation subgroup (group 2A) and the high intermediate upregulation subgroup (group 2B); and downregulation (group 1). Group 3, in which most genes were overexpressed in all samples, was composed only of CC (n=28) and the 3 CC-derived cell lines (HeLa, SiHa and CaSki). In contrast, in group 1, in which the glycolysis genes were not overexpressed, almost all controls were found (n=13, 76.5%), along with 4 of the 10 HG-CINs and a group of 15 CCs (19.7%). Group 2B, like group 3, was composed only of CC, while group 2A, more like group 1, was composed of HCE, HG-CINs and CCs (Figure 1, Supplementary Table 2). In the hierarchical grouping with the samples explored with HG-Focus, the distribution of the CCs and controls was very similar to the distribution obtained with the HG-1.0ST data (Supplementary Figure 4, Supplementary Table 2). Interestingly, the frequency of CCs with the highest clinical stage (≥ stage IIIA) predominated in group 3 (58.4%), while those with lower clinical stages (≤ stage IIB) were distributed uniformly among the 4 groups (*p=5*.*0 × 10*^*-2*^, chi-square test) (Supplementary Table 2).

**Figure 1.**
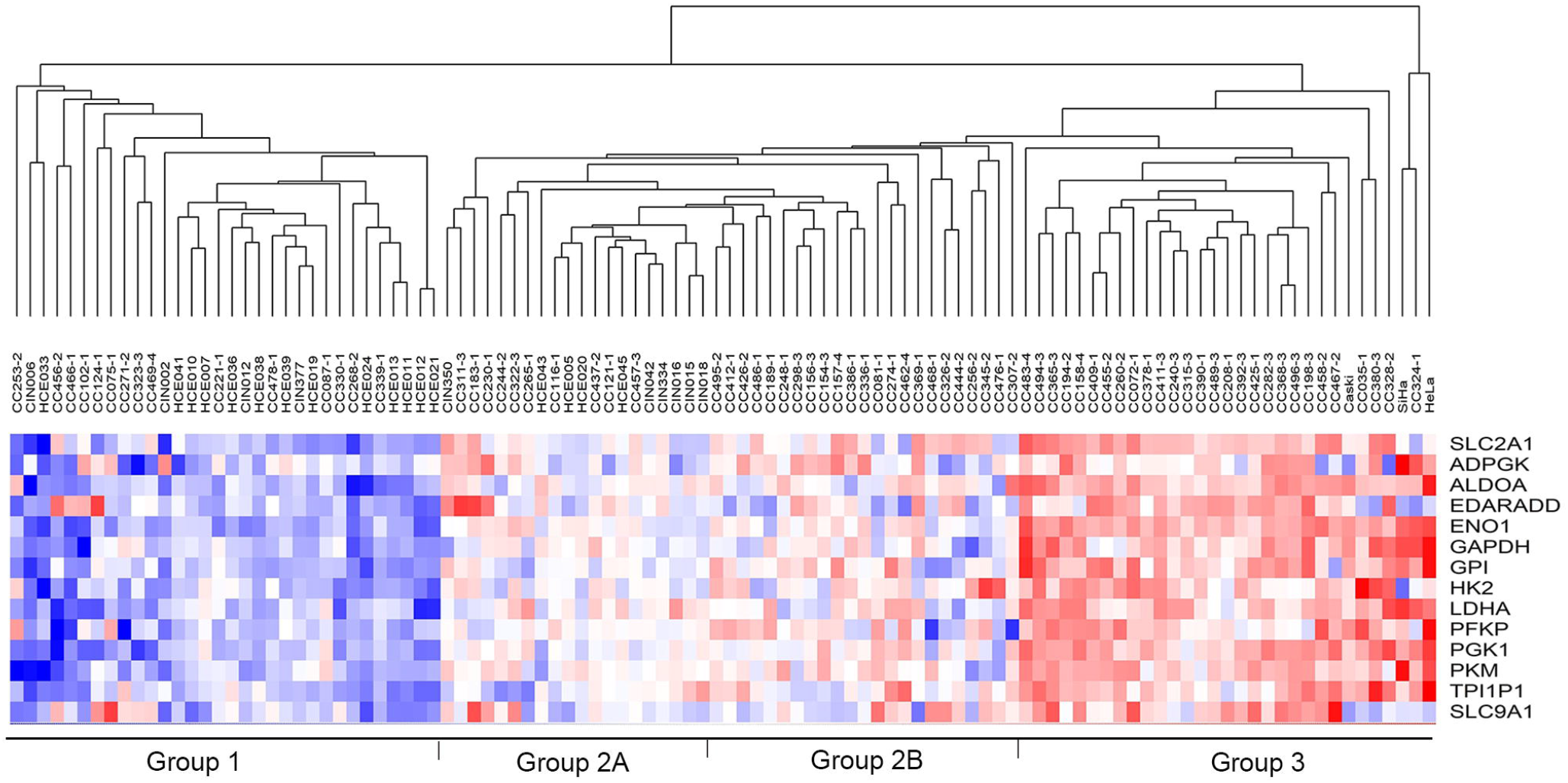
Unsupervised hierarchical cluster analysis of 14 glycolytic genes in CC. The segregation of HCE (n=17), HG-CINs (n=10) and CC (n=76) is shown according to the expression profile of 14 glycolytic genes. Four groups were distinguished: the first group with downregulation of the genes (Group 1), the second and third groups (Groups 2A and 2B) with intermediate expression levels, and the fourth group with an upregulation profile (Group 3). Each column represents a sample, and each line represents a glycolytic gene. The number at the end of the CC sample name (1 to 4) indicates the FIGO I, II, III, and IV stages of the patient. The length and subdivision of the arms represent the relationship between the samples based on the intensity of gene expression: red for upregulation, blue for downregulation, and white for no change in expression. The analysis was performed with the expression values expressed in logarithmic form (base 2).

### Effect of the gene expression of the 14 glycolysis genes on survival

Interestingly, the expression profile of the 14 genes explored significantly influences the survival of patients with CC. Almost all women with CC (95%) whose tumors did not overexpress glycolysis genes (groups 1 and 2A) survived more than 5 years. In contrast, only 55% of patients with CC who had a glycolysis overexpression profile (groups 2B and 3) survived more than 5 years (Supplementary Figure 5A), although the difference between the two groups presented marginal statistical significance (*p= 5*.*0 x10* ^*-2*^, log-rank test). To investigate which genes, contribute most significantly to that profile, survival was analyzed separately for each gene. Of the 14 genes identified with the HG-1.0ST microarray, the overexpression of only 8 of them (GAPDH, PGK1, TPI1P1, LDHA, ALDO, PFKP, ENO, and GPI) significantly reduced the % OS of the patients, with *p* values ranging from *p=1*.*0 × 10*^*-4*^ *to p= 1*.*3 × 10*^*-2*^ in the log-rank test (Supplementary Figure 5B, C, D, E, F, G, H, and I). In addition, 5 of them (GPI, PFKP, TPI1P1, GAPDH, and LDHA) were also associated with a significant reduction in % DFS (Supplementary Figure 5J-O). Of the 9 genes explored with HG-Focus, the gene expression levels of only 3 of them (ALDO, PGK1 and LDHA) were also significantly associated with the reduction in % OS (*p = 3*.*3 × 10*^*-2*^, *p = 9*.*0 × 10*^*-3*^, *and p = 5*.*0 × 10*^*-2*^, respectively, log-rank test), and only 1 (LDHA; *p = 3*.*0 × 10*^*-2*^, log-rank test) was also associated with DFS (see Supplementary Figure 6).

To determine whether the overexpression of these genes is independent of clinical stage, both variables were analyzed in a multivariate Cox proportional hazards model. Due to the small number of patients (n=61), they were grouped into two clinical groups, group 1 (≤ stage IIA, n=29) and group 2 (≥ stage IIB, n=32). Univariate analysis showed that the risk of death (hazard ratio, HR) of patients in group 2 was 3.4 (95% confidence interval (CI) = 1.1-10.4; *p= 3*.*6 × 10*^*-2*^, Cox test; Table 1) times higher than that of patients in group 1. As expected, the overexpression of 7 of the 8 genes conferred an increased risk of death, ranging from an HR of 2.8 (95% CI 1.0-7.6; *p = 3*.*7 × 10*^*-2*^) for the PGK1 gene to an HR of 9.2 (95% CI = 1.2-69.5; *p = 3*.*7 × 10*^*-2*^) for the ENO1 gene (see Table 1).

**Table 1.**
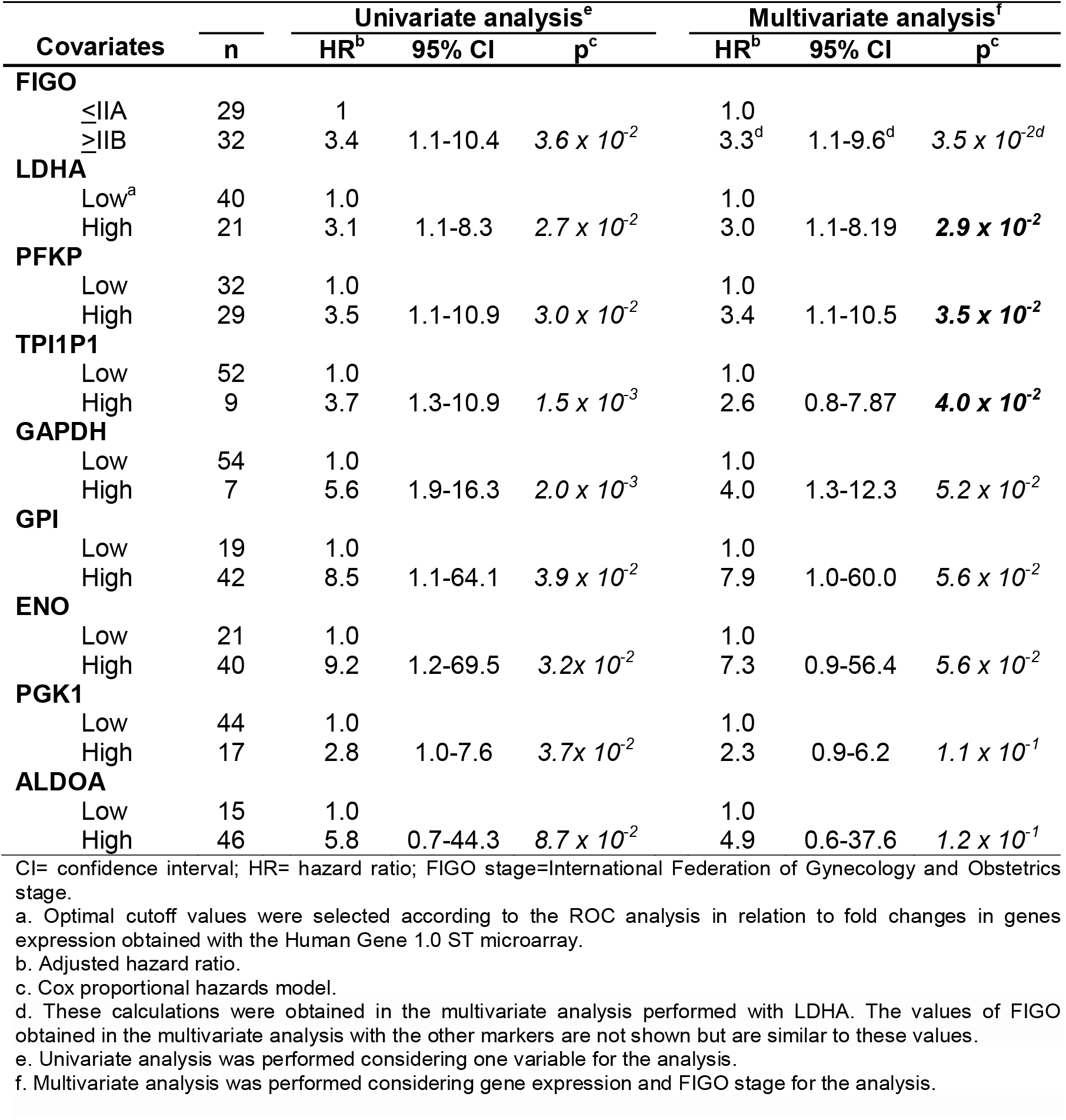
Univariate and multivariate analyses for the overall survival of patients with CC with Cox proportional hazards models including the expression of glycolytic genes explored with HG 1.0 ST microarray and FIGO clinical stage.

However, when explored in conjunction with FIGO stage in multivariate analysis, only the LDHA gene with an HR of 3.0 (95% CI =1.1-8.2; *p=2*.*9 × 10*^*-2*^), the PFKP gene with an HR of 3.4 (95% CI = 1.1-10.5; *p = 3*.*5 × 10*^*-2*^) and the pseudogene TPI1P1 with an HR of 2.6 (95% CI = 1.0-7.9; *p = 4*.*0 × 10*^*-2*^) conferred an increased risk of death, regardless of the FIGO clinical stage (Table 1). When DFS was analyzed, only LDHA with an HR of 2.7 (95% CI = 1.1-6.2; *p = 2*.*9 × 10*^*-2*^) was independent of FIGO stage (Supplementary Table 3). LDHA experimental findings were also confirmed with the HG-Focus data (see Supplementary Tables 4 and 5).

### Validation of LDHA and PFKP expression at the mRNA and protein levels

The gene expression of the LDHA and PFKP genes was validated by qRT–PCR, WB, and IH. qRT–PCR confirmed that the expression of both genes was higher in the invading CCs (n=58) than in the HCE (n=19) However, the difference was much greater for the LDHA (FC=100.3; *p=9*.*8 × 10*^*-8*^, Mann–Whitney test) than for PFKP gene (FC=4.3, *p=2*.*0 × 10*^*-6*^, Mann–Whitney test, Figure 2A). Interestingly, we found that the expression levels of LDHA and PFKP were in average 1.6 and 1.7 times higher, respectively, in patients who died and/or presented active disease (recurrence) than in those who survived and were cured (*p=9*.*0 × 10*^*-3*^ to *p=2*.*8 × 10*^*-2*^; Mann–Whitney test, see Supplementary Table 6).

**Figure 2.**
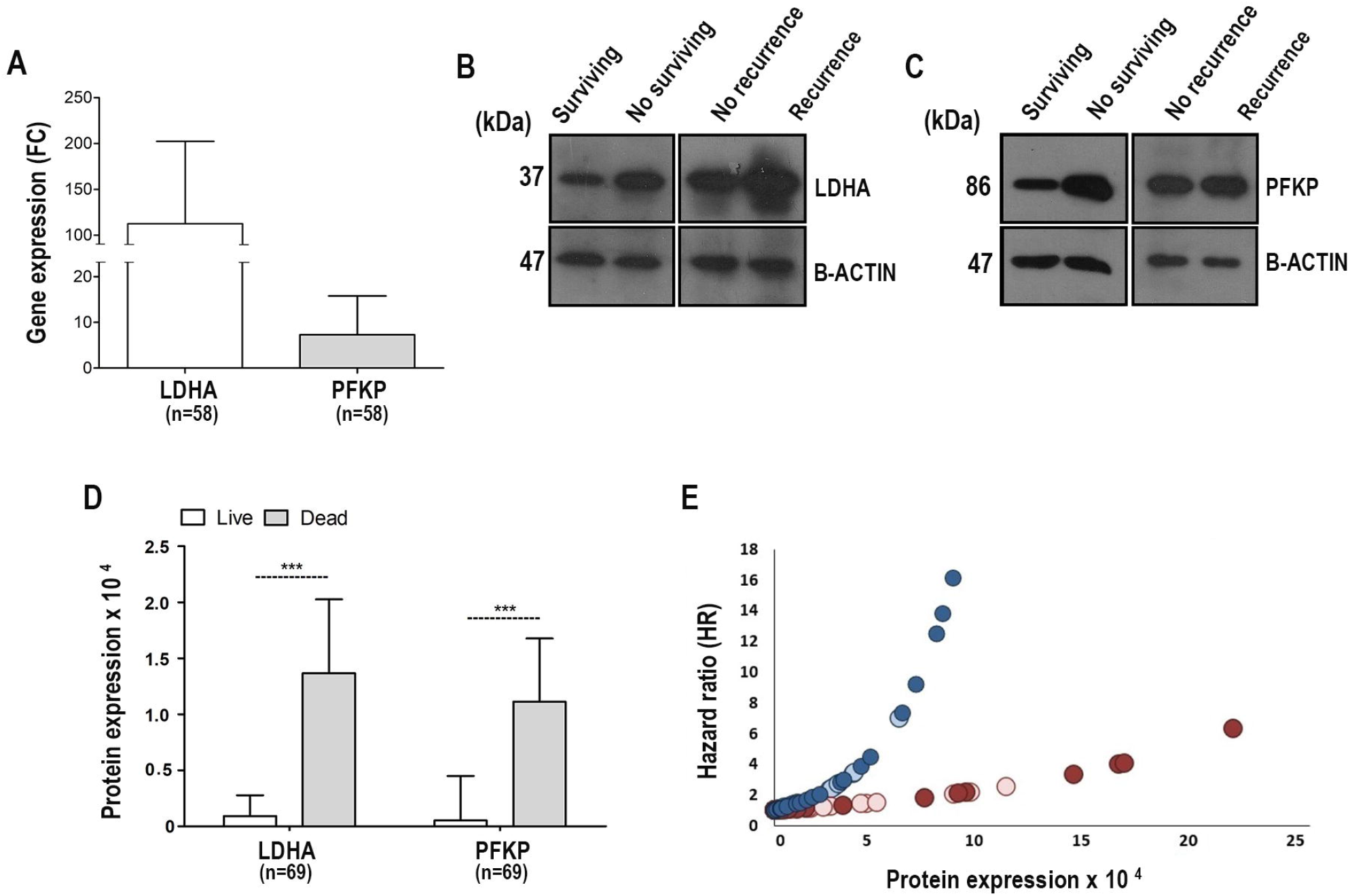
Validation of LDHA and PFKP expression in CC. The expression of LDHA and PFKP in CC was validated at the mRNA and protein levels by qRT–PCR and WB, respectively. Panel A shows the qRT–PCR analysis of LDHA and PFKP mRNA in 58 CCs. The expression was normalized with respect to the internal control (RPS13) and the control group by the double delta method using the final formula (2 ^-ΔΔ CT^). Panels B and C show representative images of WB of LDHA and PFKP expression, respectively: surviving patients, nonsurviving patients, no recurrence patients and recurrence patients. The molecular weight of the proteins is shown in kilodaltons (kDa). The protein β-actin was used as an internal control. Panel D show the mean expression + SD of LDHA and PFKP proteins between patients with CC who survived (white bars, n=47) vs. those who died (gray bars, n=22). The intensity of LDHA and PFKP was normalized with respect to β-actin. The expression is shown as optical density (OD) units. The significant differences between the groups were calculated with the Mann–Whitney U test, and *p < 0*.*05* was considered statistically significant. Panel E shows the hazard ratio (HR) analysis in relation to LDHA and PFKP protein expression in CC. The risk of dying from CC increases exponentially as protein expression (OD) increases, but it is more evident with the expression of LDHA (dark blue circles represent nonsurviving patients, while light blue circles represent surviving patients) than PFKP (dark red circles represent nonsurviving patients, while light red circles represent surviving patients).SD= standard deviation.

In addition, we confirmed the presence of the LDHA and PFKP proteins by WB in 69 CC samples (15 of the 58 tumors explored for RNAs by qRT-PCR and 54 new CCs). LDHA and PFKP proteins were expressed at higher levels in the tumors of patients who did not survive (FC=14.9, *p=3*.*0 × 10*^*-3*^ and FC=21.4, *p=1*.*8 × 10*^*-3*^, respectively; Mann–Whitney test) or who survived with the disease (FC=29.1, *p= 1*.*4 × 10-*^*4*^ and FC=17.2, *p=1*.*7 × 10* ^*-3*^, respectively; Mann–Whitney test, Figure 2B-D) compared to tumors of patients who survived or disease-free survival more than 5 years.

Additionally, we analyzed the expression of these glycolytic enzymes by IH in HCE tissues (n=12), CC tissues (n=18) and metastatic tissues (n=6) preserved in paraffin from a new group of patients. LDHA and PFKP expression levels were significantly higher in tumor tissues than in HCE tissues (FC=4.3 for LDHA and FC=27.2 for PFKP); interestingly, the expression levels of both proteins were even higher in cervical metastases than in HCE (FC= 10.4 and 42.7, respectively) see Supplementary Table 7). This suggests that overexpression of LDHA and PFKP could be an important factor not only for tumor progression but also for the development of metastases. Interestingly, we reconfirmed that LDHA expression was higher in patients who had died and/or had active disease than in those who were cured or survived more than 5 years (see Figure 3A-E and 3K). In contrast, there were not statistically significant differences in the PFKP expression between the groups (See Figure 3F-3J).

**Figure 3.**
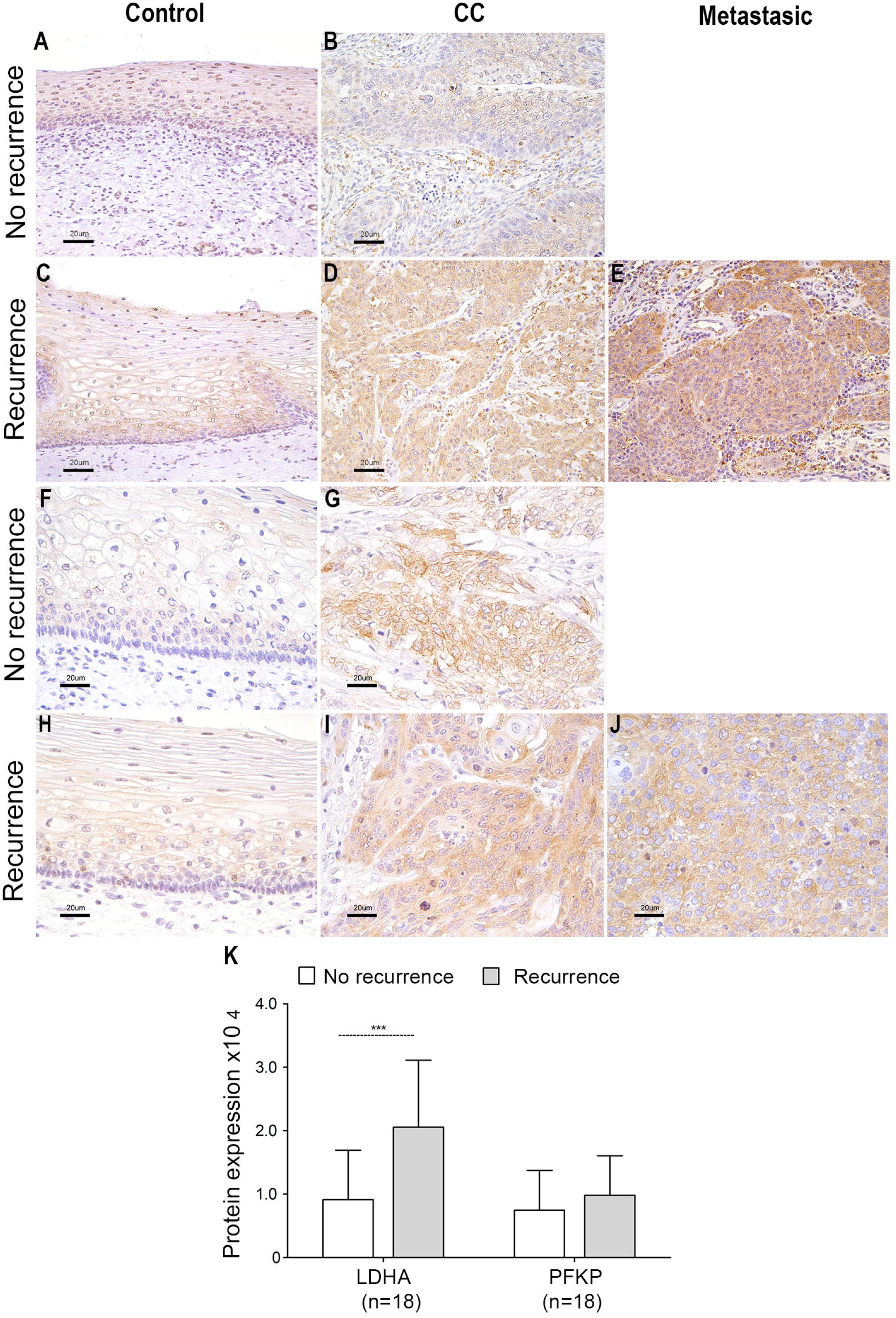
Expression of LDHA and PFKP proteins by IH. The expression of the LDHA and PFKP proteins was determined by immunohistochemistry (IH) and compared between patients who survived without the disease (No recurrence) and patients who died and/or remained with the disease (Recurrence). Histological analysis included 12 controls, 18 CC and 6 metastatic tissues. Panels A-E show the detection of the LDHA protein, and panels F-J show the detection of the PFKP protein with a specific antibody. A representative image of the experiments is shown. The specific signal for proteins is shown in brown color and counterstained with hematoxylin in violet color. Original amplification 400x; the bars measure 20 μm. Panels K show the quantitative analysis of LDHA and PFKP expression in CC tissues of patients No recurrence vs recurrence determined by IH. The average optical density and staining area of LDHA and PFKP (DOI) in the tissues were considered. The mean ± SD of three independent experiments is shown. Mann–Whitney test was performed to assess the difference between the groups, *and p < 0*.*05* was considered statistically significant. SD= standard deviation.

### The LDHA and PFKP genes, at the mRNA or protein level, are good markers of survival in CC

At both the mRNA (n=58 CC) and protein (n=69) levels, we confirmed that the overexpression of LDHA and PFKP caused a significant decrease in OS and DFS during more than 5 years of follow-up; however, the results were stronger with the analysis of proteins. The analysis of mRNA is shown in Supplementary Table 8 and Figure 4A-C and 4G-I.

**Figure 4.**
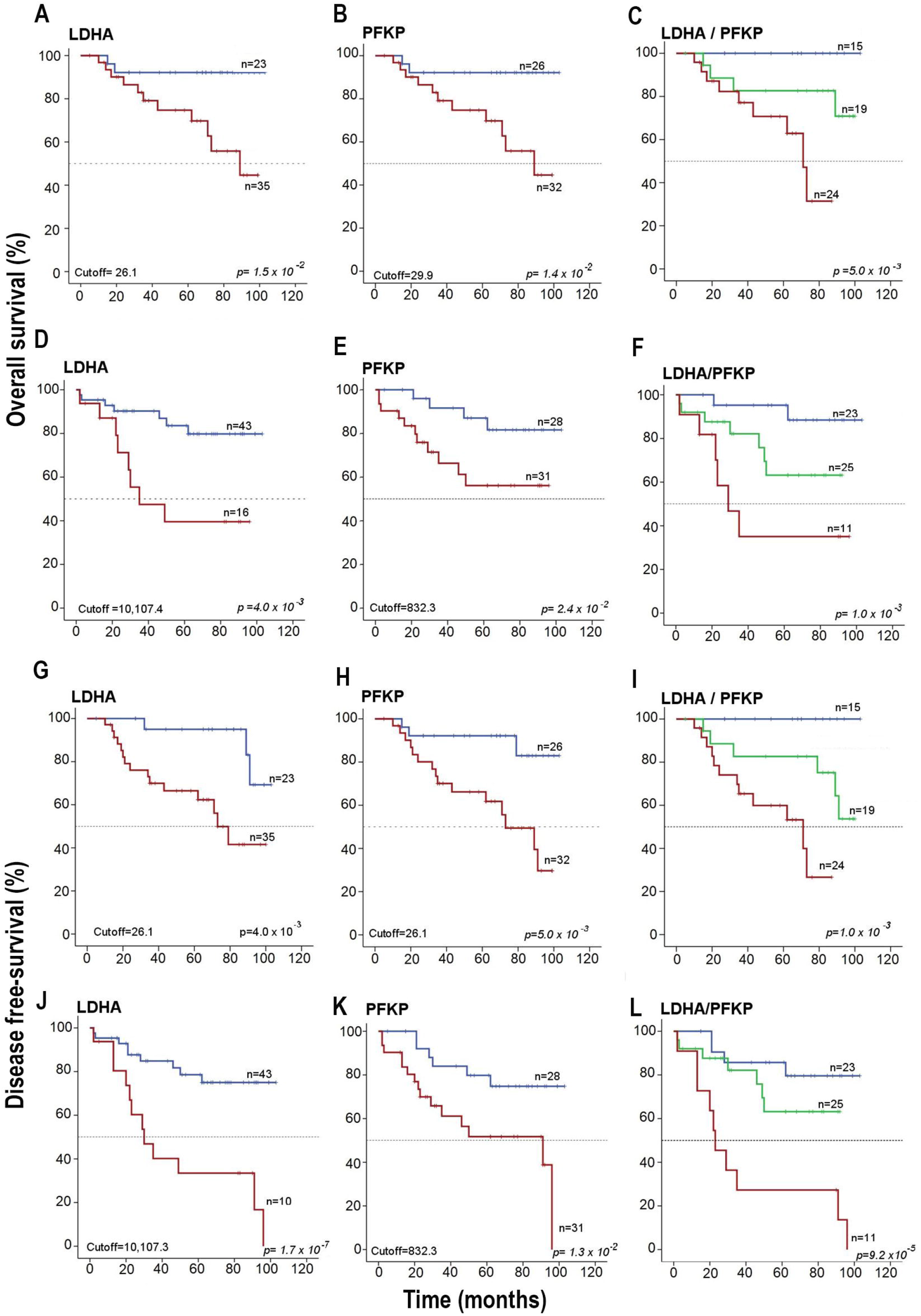
Kaplan–Meier survival curves of LDHA and PFKP. OS analysis and according to the expression of LDHA and PFKP by qRT–PCR (panels A-C) and WB (panels D-F). DFS analysis according to the expression of LDHA and PFKP by qRT– PCR (panels G-I) and WB (panels J-L). The cutoff values were calculated using ROC curves. In the overall survival analysis, the red lines include the values of nonsurviving patients, while the blue line includes the values of surviving patients. In the disease-free survival analysis, the red lines contain the values of nonsurviving patients and surviving patients with the disease, while the blue lines include the values of surviving cured patients. Censored patients are shown marked with vertical bars. The p value was calculated with the log-rank test.

At the protein level, we found that the percentage of patients who survived decreased markedly when LDHA or PFKP proteins were overexpressed compared to the group in which these proteins were not overexpressed: 39% vs. 82% and 55% vs. 83% (both *p<0*.*05*, log-rank test), respectively (see Figure 4D and 4E). Similar results were found when DFS was analyzed (Figure 4J and 4K). Interestingly, when both proteins were overexpressed (LDHA+/PFKP+), OS and DFS decreased dramatically to 29% and 23%, respectively; in contrast, when there was a single overexpressed protein, the survival rate was 64% in patients with and without the active disease, and when neither of these two proteins was expressed, the survival rate was 90% (*p=1*.*0 × 10*^*-3*^ and *p= 9*.*2 × 10*^*-5*^, log-rank test; see Figure 4F and 4L).

With the univariate Cox analysis, the risk of dying was much higher with advanced FIGO stages than with the overexpression of each of the two markers (Table 2). However, when both markers were overexpressed, they confer a greater risk of death than FIGO [HR =7 (95% CI 1.6-31.1, p=1.0 × 10 ^-2^) versus HR=8.1 (95% CI=2.6-26.1; p=4.3 × 10^−4)^]. Similar figures were seen for DFS (Table 2). Interestingly, in the multivariate analysis, both genes remained, together with clinical stage, in the models of OS or DFS, indicating that they confer a risk of death independent of FIGO stage, even of similar magnitude or greater than that conferred by FIGO stage when both markers are overexpressed [OS:HR= 6.1 (95% CI=1.3-31.2; *p=1*.*8 × 10*^*-2*^) vs. HR= 6.6 (95% CI=1.3-32.1; *p=2*.*5 × 10*^*-2*^) and [DFS:HR= 4.8 (95% CI=1.3-17.8; p=1.8 × 10^−2^) vs. HR=5.1 (95% CI=1.5-16.6; *p=7*.*0 × 10*^*-3*^)]. In fact, the HR increased exponentially as the level of expression of these markers increased, especially that of LDHA (Figure 2E). In 5 patients, the HR was well above the average HR of 4, reaching an HR value of 12.6 in the patient with an LDHA intensity of 83,538 OD units.

**Table 2.**
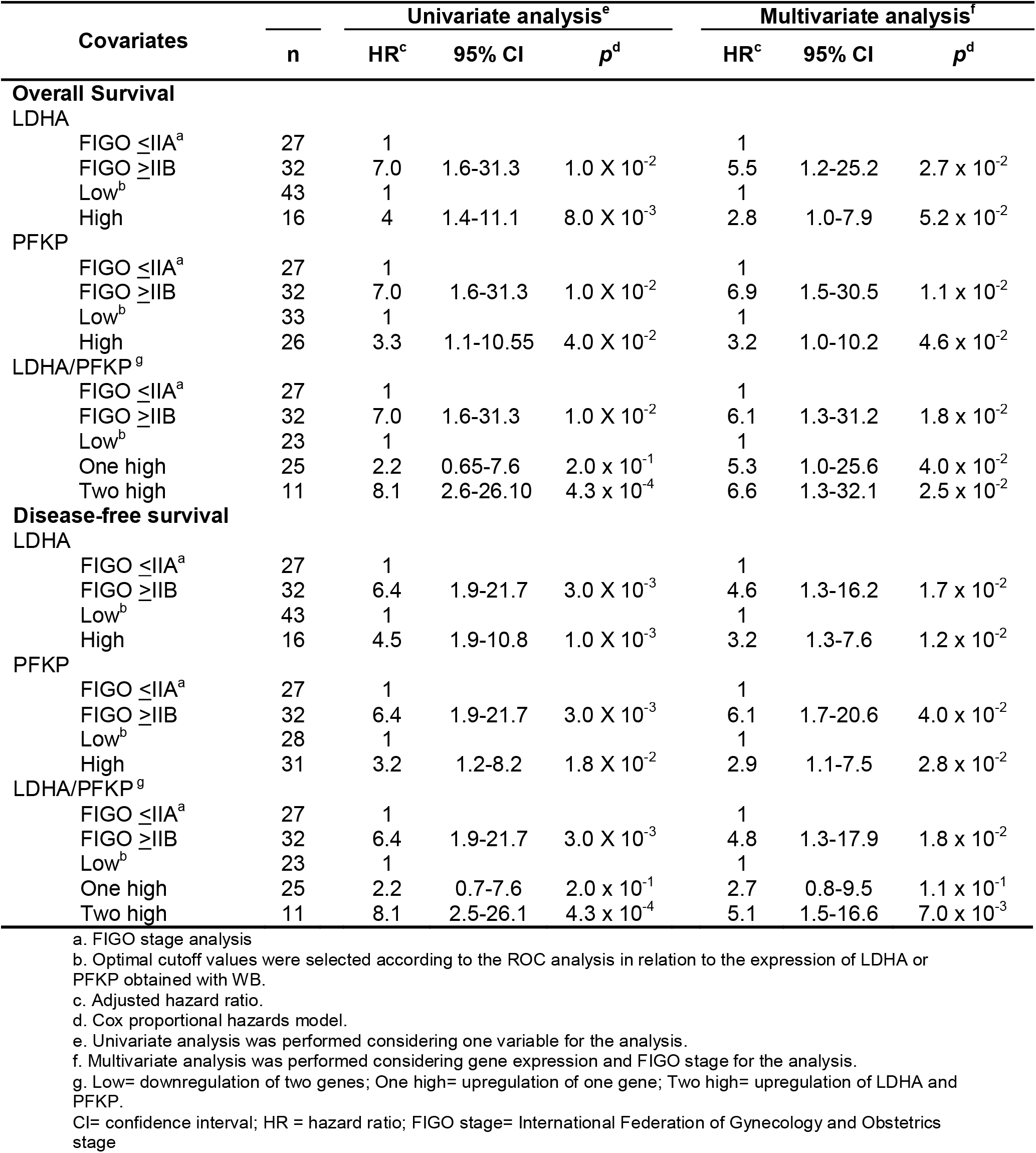
Hazard ratio analyses of patients with CC with Cox proportional hazards models including the expression of the glycolytic proteins LDHA and PFKP and FIGO clinical stage

## 4. Discussion

This is the first study in which it was identified that the overexpression of the LDHA and PFKP genes of the glycolysis pathway, both at the mRNA and protein levels, are good prognostic markers for OS and DFS in patients with CC, independent of FIGO stage. In fact, the risk of death when these two markers are elevated is equal to or greater than that of FIGO stage and increases exponentially with the protein level in the tumor, especially LDHA.

LDHA is part of the enzyme lactate dehydrogenase (LDH), which converts pyruvate into lactate. This enzyme is composed of 4 subunits, which can be A (LDHA), B (LDHB) or a combination of both [22]. Previous studies have shown that the isoforms in which the A subunit predominates favor the conversion of pyruvate into lactate, which stimulates glycolysis instead of oxidative phosphorylation. In contrast, when the B subunit predominates, the reverse happens: lactate is converted to pyruvate and metabolized by the Krebs/oxidative phosphorylation cycle [23]. In this work, we show that in CC, subunit A is overexpressed, which indicates that LDHA favors the production of lactate and, with it, anaerobic metabolism, which can provide growth advantages to CC. On the other hand, PFKP, an isoform of the enzyme phosphofructokinase 1 (PFK-1), stimulates the activity of glycolysis by catalyzing the formation of fructose 1,6-bisphosphate from fructose 6-phosphate, the first rate-limiting step of glycolysis, and consequently the production of pyruvate. The simultaneous overexpression of PFKP and LDHA makes sense for the tumor, because the concerted action of the two enzymes in CC could rapidly metabolize pyruvate to lactate, producing an acceleration of glycolysis (10-100 times faster than total glucose oxidation in the mitochondria), achieving a large amount of anaerobic ATP. This favors tumor growth and the development of more aggressive invasive tumors.

No studies have assessed the influence of LDHA gene expression on the aggressiveness and survival of patients with CC. A couple of studies focused on LDHA as a part of the tumor gene expression profile associated with metastases [24] and resistance to chemotherapy [25], however, with contradictory results. In the profile associated with resistance to chemotherapy, the LDHA gene was upregulated, while in the profile associated with tumor metastases, the LDHA gene was downregulated. Interestingly, in this last study, tumors FIGO stage ≤IIB predominated, while in the first study, tumors FIGO stage ≥IIB predominated. This could suggest that in tumors of greater clinical stage and larger size in which hypoxia probably already exists, anaerobic metabolism predominates, while in early stages with smaller tumor sizes, aerobic metabolism predominates.

On the other hand, several studies have shown that increased serum LDH activity in patients with CC was associated with poor prognosis and decreased OS [26] and DFS [27], with an increased risk of death or recurrence, independent of other clinical factors [28]. However, the limitation of these studies was that they did not demonstrate whether the quantified LDH levels came specifically from CC or from other tissues, since this enzyme is produced in several tissues.

In several types of tumors (such as squamous cell carcinoma of the skin and melanoma), increased expression of glycolysis genes is associated with increased tumor progression and decreased survival time in patients [29]. In addition, in many types of cancers, it has been observed through PET using fluorodeoxyglucose (FDG) that increased tumor glucose consumption is related to tumor aggressiveness [30]. This phenomenon has also been demonstrated in animal models. For example, in two mouse models of triple-negative breast cancer (TNBC), 4T1 and Py8119, inhibition of glycolysis resulted in reduced tumor growth and metastases, which prolonged mouse survival [31]. In CC cell lines, LDHA silencing has been shown to decrease some neoplastic features *in vitro*. For example, HeLa and SiHa cells decreased colony formation and invasion capacity when the gene is silenced by miR-34a. Interesting, when the activity of miRNA was finished, the activity of LDHA was reactivated at baseline levels favoring cell proliferation and invasion, demonstrating the importance of the expression of this gene for the tumor neoplastic phenotype [32].

Considering the importance of the neoplastic phenotype and tumor metabolism, LDHA could be a promising therapeutic target in CC. Several pharmacological inhibitors for LDHA have previously been reported for use in cancer, and there are currently several studies looking for more selective inhibitors [33] [34]. One of these compounds, gossypol, is being used in clinical trials for the treatment of malignant glioma (NCT00540722 and NCT00390403).

Although there are no PFKP reports in CC, this enzyme has been found to be overexpressed in HeLa cells [35] and related to the activation of tumor survival pathways via P44/42 mitogen-activated protein kinase (MAPK) [36]. Increased PFKP expression and activity are related to neoplastic activity, metastasis production, and decreased survival in several types of cancer, primarily brain, kidney, and breast cancers [37], [38]. Other studies have shown that the inhibition of PFKP with specific siRNAs in lung cancer cell lines [37] and murine tumor models of leukemia [39] decreased the expression of the enzyme and glycolysis, glucose, lactic acid and ATP concentrations in the supernatant of cell cultures, as well as tumor growth and progression.

Simultaneous overexpression of PFKP and LDHA has previously been described in breast cancer cell lines (MDA-MB-231), in which PFKP regulation also regulates lactate production. Interestingly, quercetin treatment impaired PFKP-LDHA signaling axis thereby inhibiting aerobic glycolysis and migration and cell invasion *in vitro* by 80% [38], demonstrating that inhibition of both enzymes may be useful in the treatment of cancers in which these enzymes are activated, as in CC.

## 5. Conclusions

The overexpression of the glycolytic enzymes Lactate dehydrogenase A (LDHA) and phosphofructokinase platelet (PFKP) was associated with poor overall and disease-free survival in CC. Overexpression of LDHA and PFKP genes increased the risk of death from CC by 8 times, and this effect was independent of the FIGO clinical stage. In fact, the risk of death from CC increased exponentially as the expression level of these markers, mainly LDHA, increased. The measurement of gene expression of these two markers could be very useful to evaluate the clinical evolution and the risk of death from CC and to make better therapeutic decisions at the beginning of treatment.

## Data Availability

All data produced in the present work are contained in the manuscript.

https://www.ncbi.nlm.nih.gov/geo/query/acc.cgi?acc=GSE52904; https://www.ncbi.nlm.nih.gov/geo/query/acc.cgi?acc=GSE39001

## 6. Abbreviations

ADPGK: ADP-dependent glucokinase
ALDOA: aldolase
AUC: area under the curve
CC: cervical cancer
DFS: disease-free survival
EDARADD: EDAR-associated death domain
ENO1: enolase 1
FIGO: International Federation of Gynecology and Obstetrics
FC: fold change
GPI: glucose-6-phosphate isomerase
GAPDH: glyceraldehyde-3-phosphate dehydrogenase
HG-CIN: high-grade cervical intraepithelial neoplasia
HK2: hexokinase 2
HR: hazard ratio
IH: immunohistochemistry
LDHA: lactate dehydrogenase A
OS: overall survival
PFKP: phosphofructokinase platelet
PGK1: phosphoglycerate kinase
PKM: pyruvate kinase M
qRT–PCR: quantitative real-time-polymerase chain reaction
SLC2A1: solute carrier family 2 member 1
SLC9A1: solute carrier family 9 member A1
TP1P1: triosephosphate isomerase 1 pseudogene 1
WB: Western blotting

## Declarations

### 7. Ethics approval and consent to participate

The study protocol was approved by the Scientific and Ethics Committee of the General Hospital of Mexico (HGM) (approval number DIC/03/311/04/051). This study, all experiments and analyses were performed in accordance with the Declaration of Helsinki. Written informed consent was obtained from all participants before their inclusion in the study.

### 8. Consent for publication

Not applicable.

### 9. Availability of data and materials

The datasets GSE52904 and GSE39001 analyzed during the current study are available in the GEO database (https://www.ncbi.nlm.nih.gov/geo/) and are available in the NCBI-GEO repository. https://www.ncbi.nlm.nih.gov/geo/query/acc.cgi?acc=GSE52904 https://www.ncbi.nlm.nih.gov/geo/query/acc.cgi?acc=GSE39001

### 12. Author contributions

Study concept and design: J.B. and V.B.S. Performed the experiments: V.B.S., A.G.C., A.M.E., I.M.M., A.A., and E.J. Analyzed the data and wrote the manuscript: V.B.S and J.B. Contributed reagents/materials: O.F.H., N.V.S. Analyzed and interpreted the image data: M.G.Z. Clinically evaluated CC patients: E.R.B., M.E.S.N., S.B.M. Provided cervical specimens from these patients: E.R.B., S.B.M. All authors read and approved the manuscript. All authors gave final approval of the version to be published.

## 13. Acknowledgments

This study was performed as part of the requirements needed to obtain the Ph.D. degree of VB from Programa de Posgrado en Ciencias Biológicas, Universidad Nacional Autónoma de México (UNAM) and received a fellowship from CONACYT (487890). We thank Dr. Valeria Barrón and Biologist Anabell Alvarado for training in some laboratory techniques.

## Figure legends

**Supplementary Figure 1. Analysis workflow of 188 cervical cancer cases**. Figure shows the analysis workflow of the 188 CC cases explored in this study. The expression of 14 genes involved in glycolysis were investigated in 76 CC with HG-1.0ST microarray and 9 genes were explored in 42 CC with HG-Focus, with 21 CCs in common. Five-year survival was investigated in 61 of 76 patients with CC explored with HG 1.0 ST and 36 of 42 patients investigated with HG-Focus. The expression of the LDHA and PFKP was validated by qRT–PCR, WB and IH. All patients explored by qRT–PCR (n=58) and 59 of 69 patients studied by WB were included in the follow-up studies, with 15 CCs in common. The protein expression of LDHA and PFKP was determined in 18 CC tissues by IH.

**Supplementary Figure 2. Expression of glycolytic genes in CC. Box plots of the expression of 14 genes of the glycolytic pathway obtained with the HG-1.0ST microarray**. The analysis was performed on 17 controls, 10 HG-CIN patients and 76 CC patients. The graphs show the value of the normalized fluorescence intensity (log 2) for each gene. The upper and lower limits of the boxes represent the 75^th^ and 25^th^ percentiles, respectively. The mean is shown as the center black line inside the boxes, and the median is shown as “+”. The whiskers represent the maximum and minimum values that lie within 1.5 times the interquartile range from the ends of the frame. Values outside this range are displayed as black dots. The *Mann–Whitney U test* was used to determine the significant differences between the groups, *\*\*p< 0*.*05, *** p< 0*.*005*.

**Supplementary Figure 3. Box plots of the expression of 9 genes of the glycolytic pathway obtained with the HG-Focus microarray**. The analysis was performed on 12 controls and 76 CC16+ patients. The graphs show the value of the normalized fluorescence intensity (log 2) for each gene. The upper and lower limits of the boxes represent the 75^th^ and 25^th^ percentiles, respectively. The mean is shown as the center black line inside the boxes, and the median is shown as “+”. The whiskers represent the maximum and minimum values that lie within 1.5 times the interquartile range from the ends of the frame. Values outside this range are displayed as black dots. The *Mann–Whitney U test* was used to determine the significant differences between the groups, *\*\*p< 0*.*05, *** p< 0*.*005*.

**Supplementary Figure 4. Unsupervised hierarchical cluster analysis of 9 glycolytic genes in CC**. The segregation of 12 controls and 42 CC HPV16+ is shown according to the expression profile of 9 glycolytic genes explored with the HG-Focus microarray, and 4 groups were distinguished: a first group with downregulation of the genes (Group 1), the second and third groups (Groups 2A and 2B) with intermediate expression levels and the fourth group with an upregulation profile (Group 3). Each column represents a sample, and each line represents a glycolytic gene. The number at the end of the CC sample name (1 to 4) indicates the FIGO I, II, III, and IV stages of the patient. The length and subdivision of the arms represent the relationship between the samples based on the intensity of gene expression: red for upregulation, blue for downregulation, and white for no change in expression. The analysis shows gene expression in log 2.

**Supplementary Figure 5**. Kaplan–Meier survival curves of HG-1.0 ST microarray. The OS (A–I panels) and DFS (J-O panels) of patients with CC were analyzed according to the expression of the 14 glycolytic genes obtained with the HG-1.0ST microarray using the Kaplan–Meier method with the SPSS program. The cutoff values of the expression of each gene that best separated the women were calculated using ROC curves. In the overall survival analysis, the red lines include the values of nonsurviving patients, while the blue line includes the values of surviving patients. In the disease-free survival analysis, the red lines contain the values of nonsurviving patients and surviving patients with the disease, while the blue lines include the surviving cured patients. The mean follow-up time of patients 60 months after the initial diagnosis. Censored patients are shown marked with vertical bars. The *p value* was calculated with the log-rank test.

**Supplementary Figure 6. Kaplan–Meier survival curves**. The OS (A-J panels) and DFS (K panel) of patients with CC were analyzed according to the expression of the 9 glycolytic genes obtained with the HG-Focus microarray using the Kaplan–Meier method. The cutoff values of the expression of each gene that best separated the patients were calculated using ROC curves. In the overall survival analysis, the red lines include the values of nonsurviving patients, while the blue line includes the values of surviving patients. In the disease-free survival analysis, the red lines contain the values of nonsurviving patients and surviving patients with the disease, while the blue lines include the values of surviving cured patients. Censored patients are shown marked with vertical bars. The *p value* was calculated with the log-rank test.

