## Supplementary tables for "Gene expression levels of the glycolytic enzymes lactate dehydrogenase A (LDHA) and phosphofructokinase platelet (PFKP) are good predictors of survival time, recurrence and risk of death in cervical cancer"

**Supplementary Table 1.** Clinicopathological characteristics

of patients with cervical cancer

| **Patient characteristics^a^** | **Total N= 188 (%)** |
| --- | --- |
| **FIGO stage** |  |
| ≤IA2 | 3 (1.6) |
| IB1 | 56 (29.8) |
| IB2 | 26 (13.8) |
| IIA | 10 (5.3) |
| IIB | 53(28.2) |
| IIIA | 2(1.1) |
| IIIB | 26(13.8) |
| ≥IV | 12 (6.4) |
| **Tumor Histology^b^** |  |
| ACC | 33(17.6) |
| SCC | 4(2.1) |
| ASCC | 151 (80.3) |
| **Viral type** |  |
| CC16+ | 132(70.2) |
| CCHPVs^c^+ | 56 (29.8) |
| **Recurrence** |  |
| No | 130(69.1) |
| Sí | 58(30.9) |
| **Mortality** |  |
| No | 133(70.7) |
| Sí | 55(29.3) |

a. All patients received complete clinical evaluation and were treated with surgery, radiation,

chemotherapy, or a combination of these according to American Cancer Society guidelines.

b. ACC= Adenocarcinoma; SCC=Squamous Cell Carcinoma, ASCC= Adenosquamous Cell Carcinoma.
c. Positive for HPVs 11, 18, 31, 33, 35, 42, 45, 51, 52, 53, 58, 59, 64 and 68.

**Supplementary Table 2.** Segregation of controls, HG-CINs, CCs and cell lines according to glycolytic gene expression profile with the HG 1.0 ST and HG-Focus microarrays.

| **Clinical groups** | **n** | **Glycolytic gene expression** | | | | ***p value^a^*** |
| --- | --- | --- | --- | --- | --- | --- |
|  |  | Frequency: n (%) | | | |  |
|  |  | **Downregulation** | **Intermediate downregulation** | **Intermediate upregulation** | **Upregulation** |  |
|  |  | Group 1 | Group 2A | Group 2B | Group 3 |  |
| **HG 1.0 ST** |  |  |  |  |  |  |
| Control | 17 | 13 (76.5) | 4 (23.5) | 0 | 0 |  |
| HG-CIN | 10 | 4 (40.0) | 6 (60.0) | 0 | 0 | *5.8 x 10^-1b^* |
| Cell lines | 3 | 0 | 0 | 0 | 3 (100) | *3.4 x 10^-2b^* |
| CC^d^ | 76 | 15 (19.7) | 10 (13.2) | 23 (30.3) | 28 (36.8) | *1.2 x 10^-3b^* |
| ≤IIB (IB1-IIB) | 52 | 13 (25.0) | 7 (13.5) | 18 (34.6) | 14 (26.9) | *5.0 x10^-2c^* |
| ≥IIIA (IIIA-IVB) | 24 | 2 (8.3) | 3 (12.5) | 5 (20.8) | 14 (58.4) |  |
| **HG-Focus** | | | | | | |
| Control | 12 | 8(66.7) | 4(33.3) | 0 | 0 |  |
| CC HPV16+ | 42 | 5(11.9) | 7(16.7) | 9(21.4) | 21(50) | *1.0 x 10^-3b^* |

CC= cervical cancer; HG-CIN= cervical intraepithelial neoplasia.

a. Chi-square test nxn. b. The *p value* was calculated with respect to the control group. c. The *p value* was calculated with respect to the ≤IIB group d. Positive for HPVs 11, 18, 31, 33, 35, 42, 45, 51, 52, 53, 58, 59, 64 and 68.

**Supplementary Table 3**. Univariate and multivariate analyses for the disease-free survival of patients with CC with Cox proportional hazards models including the expression of glycolytic genes explored with HG 1.0 ST microarray and FIGO clinical stage.

| **Covariates** |  |  | **Univariate analysis^e^** | | |  | **Multivariate analysis^f^** | | |
| --- | --- | --- | --- | --- | --- | --- | --- | --- | --- |
|  | **n** |  | **HR^b^** | **95% CI** | **p^c^** |  | **HR^b^** | **95% CI** | **p^c^** |
| **FIGO** | | | | | | | | | |
| ≤IIA | 29 |  | 1.0 |  | |  | 1.0 |  | |
| ≥IIB | 32 |  | 3.7 | 1.3-10.0 | *1.2 X 10^-2^* |  | 3.6^d^ | 1.3-9.8^d^ | *1.3 X 10^-2d^* |
| **LDHA** | | | | | | | | | |
| Low^a^ | 40 |  | 1.0 |  | |  | 1.0 |  | |
| High | 21 |  | 2.7 | 1.1-6.3 | *2.6 X 10^-2^* |  | 2.7 | 1.1-6.2 | ***2.9 X 10^-2^*** |
| **PFKP** | | | | | | | | | |
| Low | 32 |  | 1.0 |  | |  | 1.0 |  | |
| High | 29 |  | 2.4 | 0.9-5.9 | *5.8 X 10^-2^* |  | 2.3 | 0.9-5.7 | *6.2 X 10^-2^* |
| **GAPDH** | | | | | | | | | |
| Low | 54 |  | 1.0 |  | |  | 1.0 |  | |
| High | 7 |  | 5.0 | 1.9-13.3 | *1.0 X 10^-3^* |  | 3.5 | 1.3-9.6 | *6.0 X 10^-2^* |
| **TPI1P1** | | | | | | | | | |
| Low | 52 |  | 1.0 |  | |  | 1.0 |  | |
| High | 9 |  | 3.3 | 1.3-8.6 | *1.3 X 10^-2^* |  | 2.2 | 0.8-5.9 | *1.3 X 10^-1^* |
| **GPI** | | | | | | | | | |
| Low | 19 |  | 1.0 |  | |  | 1.0 |  | |
| High | 42 |  | 3.4 | 1.0-11.64 | *4.9 X 10^-2^* |  | 3.2 | 0.9-10.7 | *6.5 X 10^-2^* |
| **ENO** | | | | | | | | | |
| Low | 21 |  | 1.0 |  | |  | 1.0 |  | |
| High | 40 |  | 2.6 | 0.8-7.8 | *8.1 X 10^-1^* |  | 1.9 | 0.6-6.1 | *2.2 X 10^-1^* |
| **PGK1** | | | | | | | | | |
| Low | 44 |  | 1.0 |  | |  | 1.0 |  | |
| High | 17 |  | 2.2 | 0.9-5.2 | *7.6 X 10^-1^* |  | 1.7 | 0.7-4.1 | *2.3 X 10^-1^* |
| **ALDOA** | | | | | | | | | |
| Low | 15 |  | 1.0 |  | |  | 1.0 |  | |
| High | 46 |  | 3.4 | 0.6-8.0 | *2.3 X 10^-1^* |  | 1.9 | 0.6-6.6 | *3.0 X 10^-1^* |

CI= confidence interval; HR= hazard ratio; FIGO stage=International Federation of Gynecology and Obstetrics stage.

a. Optimal cutoff values were selected according to the ROC analysis in relation to the fold changes in gene expression obtained with the Human Gene 1.0 ST microarray.

b. Adjusted hazard ratio.

c. Cox proportional hazards model.

d. These calculations were obtained in the multivariate analysis performed with LDHA. The values of FIGO obtained in the multivariate analysis with the other markers are not shown but are similar to these values.

e. Univariate analysis was performed considering one variable for the analysis.

f. Multivariate analysis was performed considering gene expression and FIGO stage for the analysis.

.

.

**Supplementary Table 4**. Univariate and multivariate analyses for the overall survival of patients with CC with Cox proportional hazards models including the expression of glycolytic genes explored with HG-Focus microarray and FIGO clinical stage.

| **Covariates** |  |  | **Univariate analysis^e^** | | |  | **Multivariate analysis^f^** | | |
| --- | --- | --- | --- | --- | --- | --- | --- | --- | --- |
|  | **n** |  | **HR^b^** | **95% CI** | **p^c^** |  | **HR^b^** | **95% CI** | **p^c^** |
| **FIGO** | | | | | | | | | |
| ≤IIA | 26 |  | 1.0 |  | |  | 1.0 |  | |
| ≥IIB | 10 |  | 3.1 | 1.0-9.5 | *5.0 X 10^-2^* |  | 3.0^d^ | 1.0-9.1^d^ | *5.0 X 10^-2d^* |
| **LDHA** | | | | | | | | | |
| Low^a^ | 28 |  | 1.0 |  | |  | 1.0 |  | |
| High | 8 |  | 3.6 | 0.8-16.4 | *4.2 X 10^-2^* |  | 4.0 | 0.9-19.0 | ***5.0 X 10^-2^*** |
| **PGK1** | | | | | | | | | |
| Low | 20 |  | 1.0 |  | |  | 1.0 |  | |
| High | 16 |  | 9.9 | 1.2-83.5 | *3.5 X 10^-2^* |  | 8.6 | 1.4-24.3 | *2.5 X 10^-1^* |
| **ALDO** | | | | | | | | | |
| Low | 30 |  | 1.0 |  | |  | 1.0 |  | |
| High | 6 |  | 4.5 | 0.9-20.1 | *5.3 X 10^-2^* |  | 5.5 | 1.1-27.7 | *3.7 X 10^-1^* |
| **ENO1** | | | | | | | | | |
| Low | 17 |  | 1.0 |  | |  | 1.0 |  | |
| High | 19 |  | 3.0 | 0.6-15.6 | *1.9 X 10^-1^* |  | 4.2 | 0.6-27.6 | *1.3 X 10^-1^* |
| **GAPDH** | | | | | | | | | |
| Low | 17 |  | 1.0 |  | |  | 1.0 |  | |
| High | 19 |  | 2.6 | 0.5-13.4 | *2.5 X 10^-1^* |  | 2.3 | 0.4-13.0 | *4.0 X 10^-1^* |
| **SLC2A1** | | | | | | | | | |
| Low | 30 |  | 1.0 |  | |  | 1.0 |  | |
| High | 6 |  | 2.3 | 0.5-10.1 | *2.8 X 10^-1^* |  | 2.8 | 0.6-13.6 | *2.1 X 10^-1^* |
| **PKM** | | | | | | | | | |
| Low | 11 |  | 1.0 |  | |  | 1.0 |  | |
| High | 25 |  | 2.7 | 0.4-22.4 | *3.6 X 10^-1^* |  | 2.4 | 0.3-20.6 | *4.2X 10^-1^* |
| **PFKP** | | | | | | | | | |
| Low | 11 |  | 1.0 |  | |  | 1.0 |  | |
| High | 25 |  | 0.7 | 0.1-3.9 | *7.1 X 10^-1^* |  | 0.6 | 0.1-3.6 | *6.1 X 10^-1^* |
| **HK2** | | | | | | | | | |
| Low | 6 |  | 1.0 |  | |  | 1.0 |  | |
| High | 30 |  | 2.6 | 0.1-10.1 | *4.7 X 10^-1^* |  | 0.6 | 0.0-1.6 | *9.8 X 10^-1^* |

CI= confidence interval; HR= hazard ratio; FIGO stage= International Federation of Gynecology and Obstetrics stage.

a. Optimal cutoff values were selected according to the ROC analysis in relation to the mean expression of genes obtained with the Focus microarray.

b. Adjusted hazard ratio.

c. Cox proportional hazards model

d.These calculations were obtained in the multivariate analysis performed with LDHA. The values of FIGO obtained in the multivariate analysis with the other markers are not shown but are similar to these values.

e. Univariate analysis was performed considering one variable for the analysis.

f. Multivariate analysis was performed considering gene expression and FIGO stage for the analysis.

.

**Supplementary Table 5**. Univariate and multivariate analyses for the disease-free survival of patients with CC with Cox proportional hazards models including the expression of glycolytic genes explored with HG-Focus microarray and FIGO clinical stage.

| **Covariates** |  |  | **Univariate analysis^e^** | | |  | **Multivariate analysis^f^** | | |
| --- | --- | --- | --- | --- | --- | --- | --- | --- | --- |
|  | **n** |  | **HR^b^** | **95% CI** | **p^c^** |  | **HR^b^** | **95% CI** | **p^c^** |
| **FIGO** | | | | | | | | | |
| ≤IIA | 26 |  | 1.0 |  | |  | 1.0 |  | |
| ≥IIB | 10 |  | 5.6 | 1.1-27.9 | *3.4 X 10^-2^* |  | 5.4^d^ | 1.1-26.3^d^ | *3.0 X 10^-2d^* |
| **LDHA** | | | | | | | | | |
| Low ^a^ | 28 |  | 1.0 |  | |  | 1.0 |  | |
| High | 8 |  | 3.6 | 0.9-13.7 | *5.0 X 10^-2^* |  | 4.2 | 1.1-16.6 | ***3.9 X 10^-2^*** |
| **PGK1** | | | | | | | | | |
| Low | 20 |  | 1.0 |  | |  | 1.0 |  | |
| High | 16 |  | 5.4 | 1.1-26.4 | *3.6 X 10^-2^* |  | 9.9 | 1.5-24.3 | *1.6 X 10^-1^* |
| **ALDO** | | | | | | | | | |
| Low | 30 |  | 1.0 |  | |  | 1.0 |  | |
| High | 6 |  | 2.8 | 0.7-11.35 | *1.4 X 10^-1^* |  | 3.9 | 0.9-17.7 | *7.4 X 10^-1^* |
| **ENO1** | | | | | | | | | |
| Low | 17 |  | 1.0 |  | |  | 1.0 |  | |
| High | 19 |  | 3.6 | 0.7-17.8 | *1.2 X 10^-1^* |  | 3.6 | 0.7-17.8 | *1.2 X 10^-1^* |
| **GAPDH** | | | | | | | | | |
| Low | 17 |  | 1.0 |  | |  | 1.0 |  | |
| High | 19 |  | 2.0 | 0.5-8.0 | *3.2 X 10^-1^* |  | 1.5 | 0.3-6.6 | *5.7 X 10^-1^* |
| **SLC2A1** | | | | | | | | | |
| Low | 30 |  | 1.0 |  | |  | 1.0 |  | |
| High | 6 |  | 1.5 | 0.3-7.5 | *5.8 X 10^-1^* |  | 2.8 | 0.7-11.7 | *1.5 X 10^-1^* |
| **PKM** | | | | | | | | | |
| Low | 11 |  | 1.0 |  | |  | 1.0 |  | |
| High | 25 |  | 1.2 | 0.3-1.6 | *5.8 X 10^-1^* |  | 1.2 | 0.3-6.3 | *7.8X 10^-1^* |
| **PFKP** | | | | | | | | | |
| Low | 11 |  | 1.0 |  | |  | 1.0 |  | |
| High | 25 |  | 0.6 | 0.3-7.5 | *5.3 X 10^-1^* |  | 0.6 | 0.1-2.3 | *3.7 X 10^-1^* |
| **HK2** | | | | | | | | | |
| Low | 6 |  | 1.0 |  | |  | 1.0 |  | |
| High | 30 |  | 1.5 | 0.8-11.75 | *7.1 X 10^-1^* |  | 1.3 | 0.1-10.4 | *8.1 X 10^-1^* |

CI= Confidence interval; HR= hazard ratio; FIGO stage= International Federation of Gynecology and Obstetrics stage.

a. Optimal cutoff values were selected according to the ROC analysis in relation to the mean expression of genes obtained with the Focus microarray.

b. Adjusted hazard ratio.

c. Cox proportional hazards model.

d. These calculations were obtained in the multivariate analysis performed with LDHA. The values of FIGO obtained in the multivariate analysis with the other markers are not shown but are similar to these values.

e. Univariate analysis was performed considering one variable for the analysis.

f. Multivariate analysis was performed considering gene expression and FIGO stage for the analysis.

.

**Supplementary Table 6.** Expression of LDHA and PFKP by

qRT–PCR in clinical groups (n=58)

| **Clinical groups** | **Gene expression** | | |
| --- | --- | --- | --- |
|  | **n** | **Mean (ng)^a^ ± DE** | ***p^b^*** |
| **Overall survival** | | | |
| **LDHA** | | | |
| Surviving | 45 | 37.05 ± 30.2 | *9.0 x 10^-2^* |
| Nonsurviving | 13 | 54.99 ± 41.8 |  |
| **PFKP** | | | |
| Surviving | 45 | 40.6 ± 33.6 | *3.5 x 10^-2^* |
| Nonsurviving | 13 | 66.6 ± 51.6 |  |
| **Disease-free survival** | | | |
| **LDHA** | | | |
| No recurrence | 40 | 33.46 ± 25.07 | *9.0 x 10^-3^* |
| Recurrence | 18 | 58.0 ± 43.58 |  |
| **PFKP** | | | |
| No recurrence | 40 | 38.9 ± 33.96 | *2.8 x 10^-2^* |
| Recurrence | 18 | 63.23 + 46.04 |  |

a. Mean absolute quantification with standard curve.

The expression was normalized to that of RPS13.

b. Mann–Whitney test for surviving vs. nonsurviving or no recurrence vs. recurrence.

DE=standard deviation

**Supplementary Table 7.** Expression of LDHA and PFKP by IH in cervical cancer

| **Clinical groups** | **LDHA** | | | | | |  | **PFKP** | | | |
| --- | --- | --- | --- | --- | --- | --- | --- | --- | --- | --- | --- |
|  | **n** |  | **DOI ^a^** | **DE** | **FC^b^** | **p^c^** |  | **DOI ^a^** | **DE** | **FC^b^** | **p^c^** |
| Control | 12 |  | 34,160 | 32,153 |  |  |  | 3,124 | 4,385 |  |  |
| CC | 18 |  | 147,387 | 108,499 | 4.3 | 1.2x 10^-3^ |  | 84,935 | 63,105 | 27.2 | 7.6 x 10^-3^ |
| Metastatic | 6 |  | 356,409 | 146,719 | 10.4 | 3.2x 10^-6^ |  | 133,660 | 82,216 | 42.7 | 1.7x 10^-5^ |

a. DOI=mean integrated density.

b. Fold change (FC) was calculated with the median values as follows: clinical groups/control.

c. Mann–Whitney rank sum test of control vs. CC.

CC= cervical canc

**Supplementary Table 8**. Univariate and multivariate survival analyses of patients with CC with Cox proportional hazards models including the expression of glycolytic genes and FIGO clinical stage by qRT–PCR.

| **Covariates^a^** | |  | |  | | **Univariate analysis^e^** | | | | | | | |  | **Multivariate analysis^f^** | | | | |
| --- | --- | --- | --- | --- | --- | --- | --- | --- | --- | --- | --- | --- | --- | --- | --- | --- | --- | --- | --- |
|  |  | **n** | |  | | **HR^c^** | | **95% CI** | | ***p*^d^** | | | |  | **HR^c^** | **95% CI** | ***p*^d^** | | |
| **Overall Survival** | | | | | | | | | | | | | | | | | | | |
| LDHA | | | | | | | | | | | | | | | | | | | |
| FIGO ≤IIA^a^ | | 29 | |  | | 1.0 | |  | | | | | |  | 1 |  | | | |
| FIGO ≥IIB | | 29 | |  | | 2.6 | | 0.9-8.1 | | 8.7 X 10 ^-2^ | | | |  | 1.7 | 0.5-5.5 | 1.2 X 10 ^-1^ | | |
| Low^b^ | | 23 | |  | | 1.0 | |  | | | | | |  | 1 |  | | | |
| High | | 35 | |  | | 5.6 | | 1.2-26.3 | | 2.9 X 10 ^-2^ | | | |  | 5.0 | 1.0-23.8 | 4.4 x 10 ^-2^ | | |
| PFKP | | | | | | | | | | | | | | | | | | | |
| FIGO ≤IIA | | 29 | |  | | 1 | |  | | | | | |  | 1 |  | | | |
| FIGO ≥IIB | | 29 | |  | | 2.6 | | 0.9-8.1 | | | | 8.7X 10 ^-2^ | |  | 2.0 | 0.6-6.7 | | | 4.4 x 10 ^-2^ |
| Low | | 26 | |  | | 1.0 | |  | | | | | |  | 1 |  | | | |
| High | | 32 | |  | | 5.4 | | 1.2-24.6 | | 2.8 X 10 ^-2^ | | | |  | 5.0 | 1.1-22.9 | 3.7 x 10^-2^ | | |
| LDHA/PFKP ^g^ | | | | | | | | | | | | | | | | | | | |
| FIGO ≤IIA | | 29 | |  | | 1 | |  | | | | | |  | 1 |  | | | |
| FIGO ≥IIB | | 29 | |  | | 2.6 | | 0.9-8.1 | | | | 8.7X 10 ^-2^ | |  | 2.3 | 0.8-6.6 | | | 1.3 x 10^-1^ |
| Low/one high | | 34 | |  | | 1 | |  | | | | | |  | 1 |  | | | |
| Two high | | 24 | |  | | 6.7 | | 1.7-24.4 | | 7.0 x 10^-3^ | | | |  | 6.4 | 1.7-26.6 | 7.0 x 10^-3^ | | |
|  | **Disease-free survival** | | | | | | | | | | | | | | | | | | |
|  | LDHA | | | | | | | | | | | | | | | | | | |
|  | FIGO ≤IIA | | 29 | |  | | 1 | |  | | | | |  | 1 |  | | | |
|  | FIGO ≥IIB | | 29 | |  | | 2.6 | | 0.9-7.6 | | 7.3 X 10 ^-2^ | | |  | 2.5 | 0.9-7.0 | 8.3 X 10 ^-2^ | | |
|  | Low | | 23 | |  | | 1 | |  | | | | |  | 1 |  | | | |
|  | High | | 35 | |  | | 5.5 | | 1.5-19.7 | | 9.0 X 10 ^-3^ | | |  | 4.8 | 1.3-17.3 | 1.8 x 10 ^-2^ | | |
|  | PFKP | | | | | | | | | | | | | | | | | | |
|  | FIGO ≤IIA | | 29 | |  | | 1 | |  | | | | |  | 1 |  | | | |
|  | FIGO ≥IIB | | 29 | |  | | 2.6 | | 0.9-7.6 | | | | 7.3X 10 ^-2^ |  | 2.1 | 0.8-6.0 | | 1.5 x 10 ^-2^ | |
|  | Low | | 26 | |  | | 1 | |  | | | | |  | 1 |  | | | |
|  | High | | 32 | |  | | 5.0 | | 1.4-17.2 | | 1.8 X 10 ^-2^ | | |  | 4.5 | 1.3-15.8 | 1.7 x 10^-2^ | | |
|  | LDHA/PFKP expression ^g^ | | | | | | | | | | | | | | | | | | |
|  | FIGO≤IIA | | 29 | |  | | 1 | |  | | | | |  | 1 |  | | | |
|  | FIGO≥IIB | | 29 | |  | | 2.6 | | 0.9-7.6 | | 7.3 X 10 ^-2^ | | |  | 2.6 | 0.9-7.6 | 7.3 X 10 ^-2^ | | |
|  | Low/one high | | 34 | |  | | 1 | |  | | | | |  | 1 |  | | | |
|  | Two high | | 24 | |  | | 7.7 | | 2.2-23.5 | | 1.0 x 10^-3^ | | |  | 7.1 | 2.2-27.1 | 1.0 x 10^-3^ | | |

a. FIGO stage analysis

b. Optimal cutoff values were selected according to the ROC analysis in relation to the expression of LDHA and PFKP obtained with qRT–PCR.

c. Adjusted hazard ratio.

d. Cox proportional hazards model.

e. Univariate analysis was performed considering one variable for the analysis.

f. Multivariate analysis was performed considering gene expression and FIGO stage for the analysis.

g. Low/one high= downregulation of two genes or upregulation of one gene; Two high= upregulation of LDHA and PFKP.

CI= confidence interval; HR = hazard ratio; FIGO stage= International Federation of Gynecology and Obstetrics stage
