## Supplementary text 1 for "Gene expression levels of the glycolytic enzymes lactate dehydrogenase A (LDHA) and phosphofructokinase platelet (PFKP) are good predictors of survival time, recurrence and risk of death in cervical cancer"

**Supplementary material 1.**

**2. Methods**

**DNA isolation**

The DNA was extracted using the PureLink Genomic DNA Kit (Invitrogen, Carlsbad, CA, USA).

**Detection and HPV typing**

HPV detection was performed by PCR using universal primers located in the HPV L1 gene (MY09/MY11, GP5+/6+, and L1C1), as described previously [18]. The HBB gene was used as an internal control to assess the quality of the DNA. The HPV types were identified by sequencing the amplified bands using the fluorescent cycle-sequencing method (BigDye Terminator Ready Reaction Kit; Applied Biosystems, Carlsbad, CA, USA). Sequence analysis was performed using an ABI PRISM 3130xl Genetic Analyzer system (Applied Biosystems). Each band sequenced was analyzed with the FASTA sequence similarity. The average identity percentage of HPV types detected was 98.7% (91-100%) when compared to the reference sequences.

**Glycolytic gene expression and data analysis**

The HG 1.0 ST was standardized with the Robust Multiarray Average algorithm in the Affymetrix expression console, and the HG-Focus was standardized with the Robust Multichip Average algorithm of FlexArray software [16,18].

The identification of glycolytic genes differentially expressed between CC, HG-CIN and controls was performed with the SAM algorithm (SAM version 3.0, http://statweb.stanford.edu/~tibs/SAM/) using a cutoff fold change (FC) value of ≥1.5, a general false discovery rate of 0%, and a local false discovery rate of ≤10%. The normalized intensity values (UI) were transformed to log 2 values for analysis. We identified 14 glycolytic genes that met the selection criteria: SLC2A1, ADPGK, HK2, GPI, PFKP, ALDOA, TPI1P1, GAPDH, PGK1, ENO1, PKM, LDHA, SLC9A1 and EDARADD. We performed an unsupervised hierarchical grouping analysis using dChip software (version 1.6, http://www.hsph.harvard.edu/cli/complab/dchip/) with the parameters of Euclidean metric distance, linkage average method, genes ordered by the time peak and rows standardized by the mean [16].
