## Supplementary figures and images for "Gene expression levels of the glycolytic enzymes lactate dehydrogenase A (LDHA) and phosphofructokinase platelet (PFKP) are good predictors of survival time, recurrence and risk of death in cervical cancer"

### Supplementary Figure 1

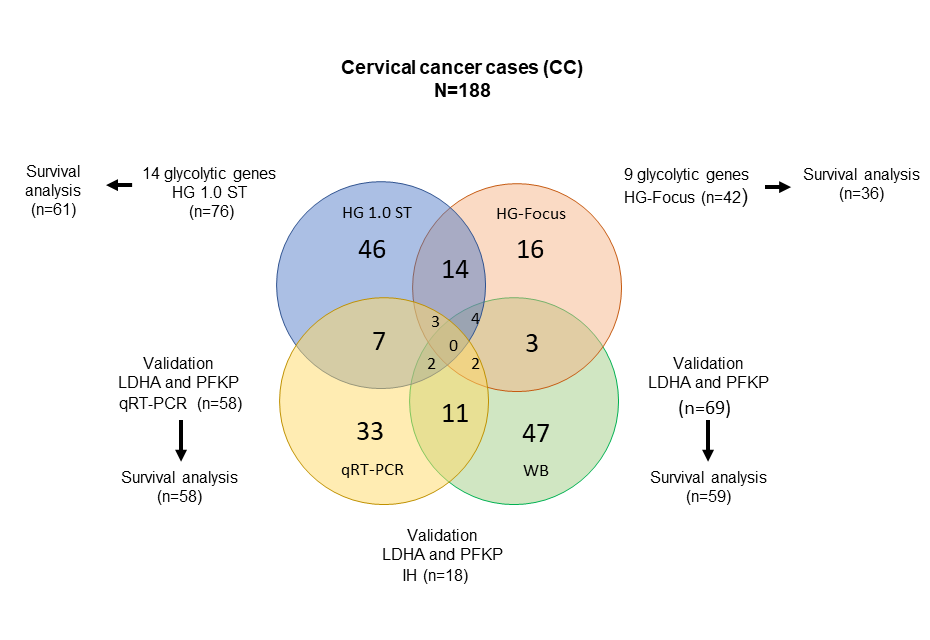

### Supplementary Figure 2

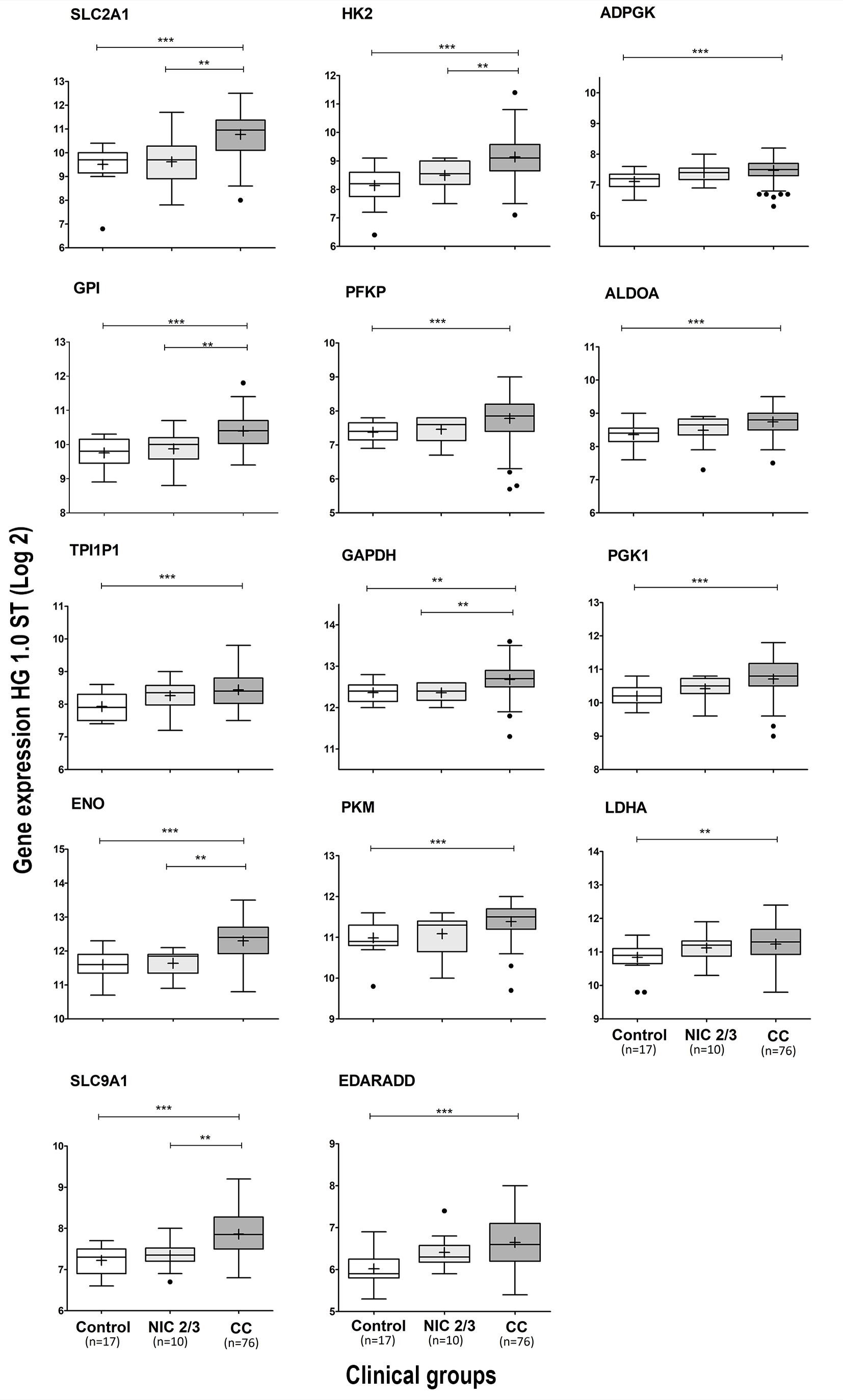

### Supplementary Figure 4

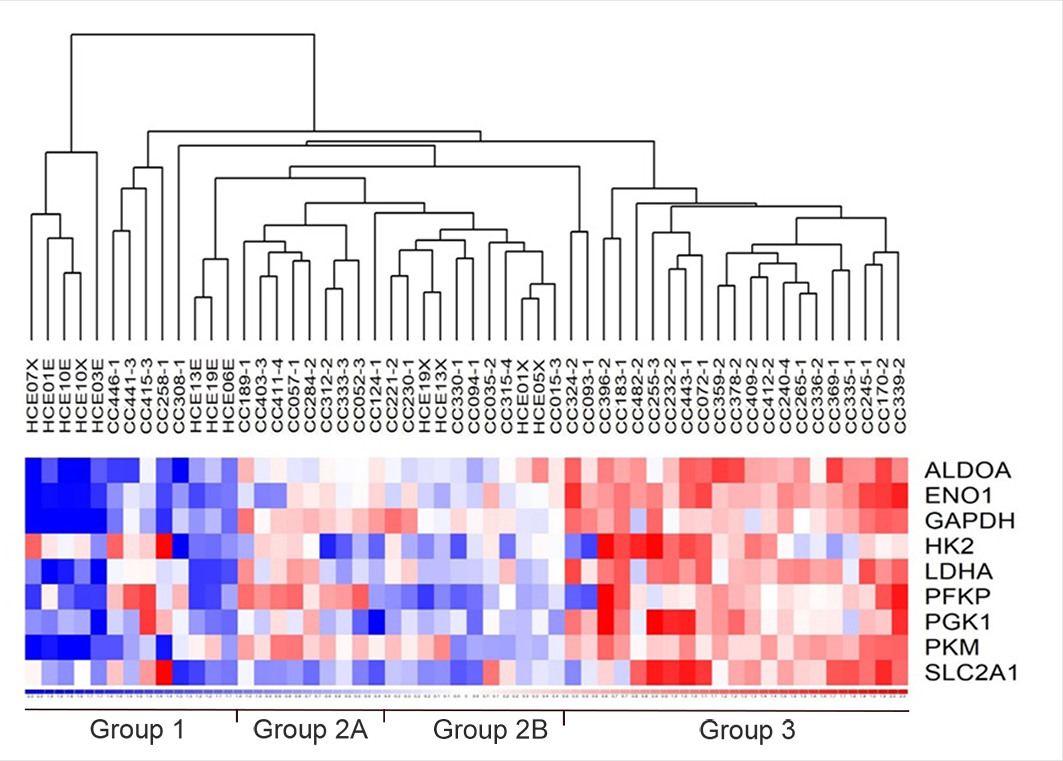

### Supplementary Figure 5

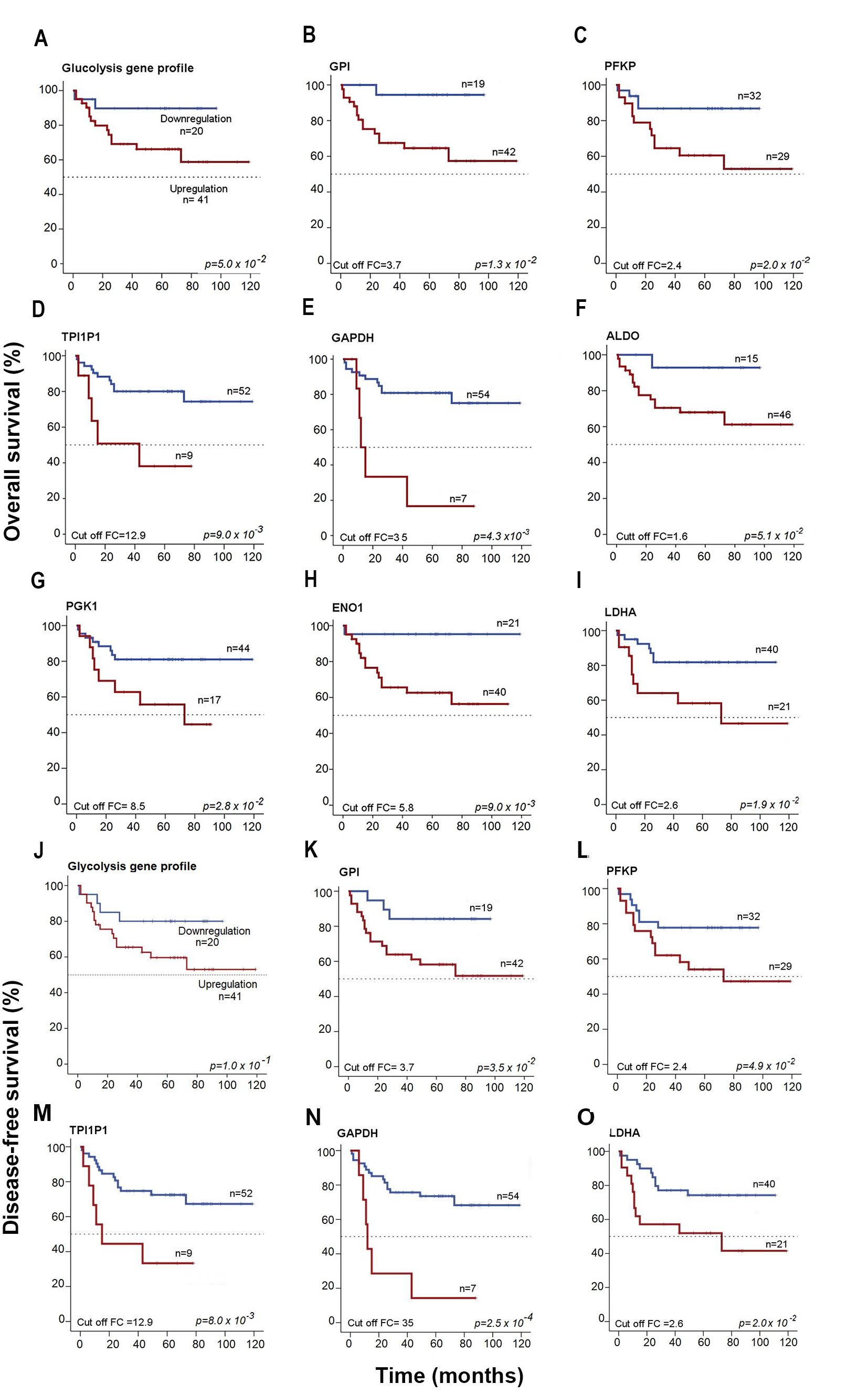

### Supplementary Figure 6

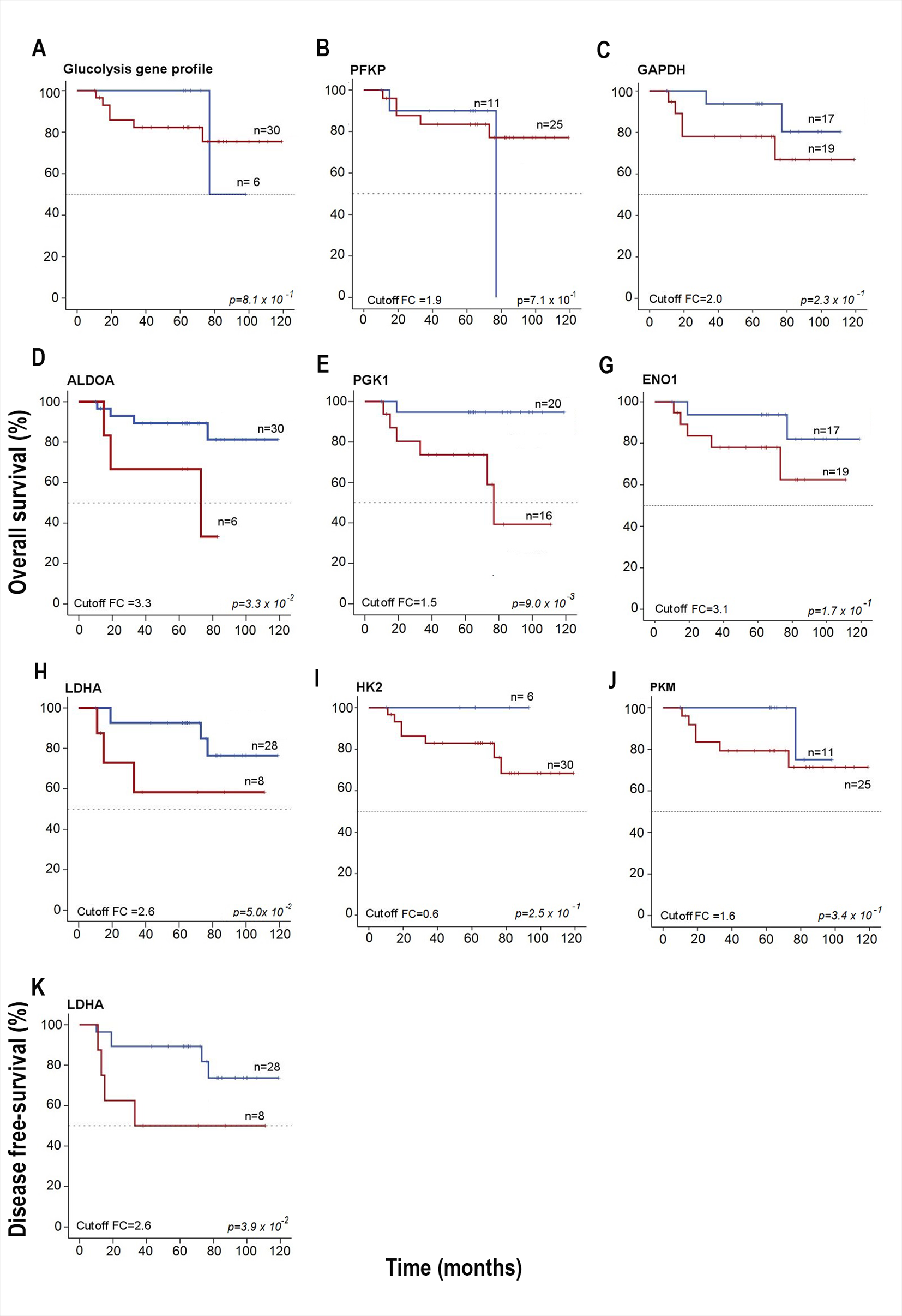
